# Misinformation can prevent the suppression of epidemics

**DOI:** 10.1101/2021.08.23.21262494

**Authors:** Andrei Sontag, Tim Rogers, Christian Yates

## Abstract

The effectiveness of non-pharmaceutical interventions, such as mask-wearing and social distancing, as control measures for pandemic disease relies upon a conscientious and well-informed public who are aware of and prepared to follow advice. Unfortunately, public health messages can be undermined by competing misinformation and conspiracy theories, spread virally through communities that are already distrustful of expert opinion. In this article, we propose and analyse a simple model of the interaction between disease spread and (mis-) information dynamics in a heterogeneous population composed of both trusting individuals who seek quality information and will take precautionary measures, and distrusting individuals who are susceptible to misinformation. We show that, as the density of the distrusting population increases, the model passes through a phase transition to a state in which major outbreaks cannot be suppressed. Our work highlights the urgent need for effective measures to combat the spread of misinformation.

## 1. Introduction

Non-pharmaceutical interventions, such as mask-wearing and social distancing, are important tools to tackle the spread of emergent pandemic disease [1]. They can be used to slow the progress of an outbreak or suppress it completely if carefully implemented and strictly adhered to [2, 3]. There are, however, substantial drawbacks, including immediate economic damage to certain industries and longer-term negative effects of social isolation on the wellbeing of individuals. Beyond any rational cost-benefit analysis, these measures also evoke a strong emotional response. The concept of a sudden government demand for people to change their behaviour and limit social interaction in the face of an unseen enemy is at odds with prevailing western liberal ideologies of the importance of individual freedoms. Unsurprisingly, these policies do not enjoy universal support and public perceptions are a key consideration in deciding implementation details – it has been widely reported that the UK government delayed action in spring 2020 for fear that “behavioural fatigue” would limit public adherence to social distancing rules [4].

Much of the effectiveness of social distancing strategies relies upon the behaviours of individuals; how conscientiously they follow advice and how successfully they encourage others to do the same. Many factors affect the degree to which individuals will change their behaviours: perception of the risk to themselves and to others; quality and availability of information; trust in the government and/or scientific establishment; the attitudes of their social contacts to these issues. Importantly, there is considerable feedback between these factors and the progression of the epidemic itself [5]. Previous modelling studies have examined the interplay between risk perception (modelled as an information spreading process), behaviour, and epidemic dynamics. Methodologies applied include: game theory models for social distancing [6]; opinion dynamics for behavioural change [7]; models of fear of infection and fear of the control [8, 9]; SIS-like models with aware categories on networks where behaviour changes according to health status and risk perception [10]; SIR models with aware and unaware states [11]; and awareness-driven reduction in contacts [12].

Of particular interest is the work by Funk et al. [13] investigating the interplay between behavioural change and disease spread through a stochastic SIR-like model [14] coupled with an awareness model in which individuals transmit information through a contact-based network. The authors characterise the dynamics of information spread by the following two event types: (1) *transmission of awareness*, occurring when an individual encounters someone with more up-to-date information, and (2) *fading of awareness*, which describes gradual relaxation back to a low-awareness state. The feedback between disease and information dynamics occurs through the generation of new information by infected individuals when they realise their condition, after which they and their contacts may adopt safer behaviours to limit further spread of the infection. In [15] (and similarly in independent work of Kiss et al. [16]) a simplified version of this model was explored with only two states of aware-unaware individuals, in which it was shown that awareness-driven behaviour change can alter the outbreak conditions and can make it impossible for a disease to establish itself in the population.

The works discussed above share common (often unstated) assumptions about the populations they describe. They model homogeneous populations of individuals that believe in the existence of the disease, are prepared to alter their behaviour to reduce transmission, and have been correctly informed how to do so. Recent evidence sadly shows the extent to which these assumptions fail [17–21].

In this study, we extend the established model of Funk et al. [13] to include a subset of individuals who are distrustful of official advice, susceptible to misinformation, and more likely to engage in risky behaviours. We show that the presence of such dynamics can radically reduce the effectiveness of epidemic intervention strategies based on behaviour change. We observe a new dynamical threshold corresponding to a phase transition between regimes of successful disease suppression and large outbreaks. A theoretical estimate of the critical parameters for this transition is obtained in the limit of fast information spread.

The article is organised as follows. In Sec. 2, we propose a model of epidemics with behaviour change feedback in a heterogeneous population composed of trusting and distrusting individuals. Results from the modelling approach are presented in Sec. 3. We discuss the observed phase transition in Sec. 3.1 and derive a theoretical estimate for the fast spread of information limit in Sec. 3.2. We conclude with a brief discussion of our results in Sec. 4.

## 2. The Model

We extend and modify the model presented in [13] with the introduction of a heterogeneous population composed of (i) *trusting* individuals who seek information of higher quality and will take precautionary measures, and (ii) *distrusting* individuals, who are susceptible to misinformation. We consider a population of fixed size *N* and SIR dynamics for disease spread [22]. In the SIR model, the population is compartmentalised into three disease states: susceptible *S*, infected *I* and recovered *R*. We distinguish subpopulations in our model by two indexes giving the behavioural group to which they belong, i.e., trusting (T) or distrusting (D), and the quality, *i*, of the information they possess. Information quality decreases with increasing index *i* so that, for example, an individual in the group *S*_*T*,0_ is a susceptible member of the trusting population and possesses the best quality information.

The two groups of trusting and distrusting individuals are distinguished by the way they obtain information. Trusting individuals of any disease state *X* ∈ {*S, I, R*} with information of quality *j, X*_*T,j*_, will accept new information when they encounter individuals of any group with information of better quality *i*, i.e., *X*_*T,j*_ + *X*_*Y,i*_ → *X*_*T,i*+1_ + *X*_*Y,i*_, where *i* < *j* and *Y* can be either trusting (T) or distrusting (D). The “+1” in the information subscript represents a loss in quality when the information is exchanged. Hence, trusting individuals possess the same information dynamics as assigned to all individuals in [13]. In contrast, distrusting individuals *X*_*D,j*_ receive information when encountering individuals with lower quality information, i.e., *X*_*D,j*_ + *X*_*Y,i*_ → *X*_*D,i*+1_ + *X*_*Y,i*_, where *i* > *j*. Parameters *α*_*T*_ and *α*_*D*_ control the encounter rates for each subpopulation.

Besides information transmission, we also assume a *fading* effect. Due to the fading effect, information loses quality if not refreshed, increasing the individual’s index by 1, i.e., *X*_*i*_ → *X*_*i*+1_. This occurs at a rate *λ*. This dynamic accounts for reluctance of the population to continue with inconvenient measures when they do not perceive immediate risk.

The described information dynamics lead to the following equations representing the changes of population size at each information level *k* for each group of trusting and distrusting individuals,

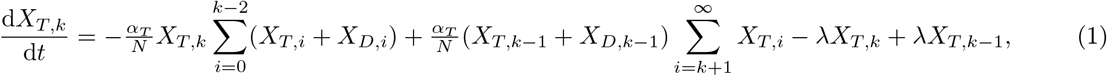

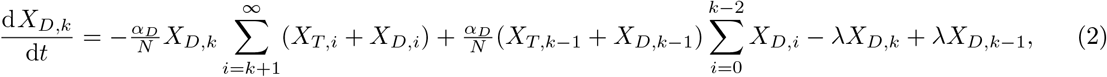

which are valid for *k* ∈ {0, 1, …} if we take *X*_*y,j*_ = 0 for *j* < 0. The first term in Eq. (1) corresponds to individuals at the information level *k* moving to a better quality of information after interacting with either trusting or distrusting better-informed individuals. The second term corresponds to trusting individuals with worse information that interact with individuals at the level *k* − 1, acquiring information of quality *k*. The last two terms describe the changes due to the fading effect. Similarly, the first term in Eq. (2) gives the changes due to distrusting individuals with information quality *k* obtaining worse quality of information. The second term is the gain due to individuals with better information quality receiving information from level *k* − 1. The final terms again describe the changes due to information fading. Once some level of disease awareness is present in the population, the changes in distribution of the population throughout the awareness levels will follow the dynamics given by Eqs. (1) and (2).

Denoting *I*_*i*_ = *I*_*T,i*_ + *I*_*D,i*_, *t*_*i*_ = *S*_*T,i*_ + *I*_*T,i*_ + *R*_*T,i*_, and *d*_*i*_ = *S*_*D,i*_ + *I*_*D,i*_ + *R*_*D,i*_, we obtain the following set of ordinary differential equations to describe the changes in the subpopulations due to disease and behaviour dynamics,

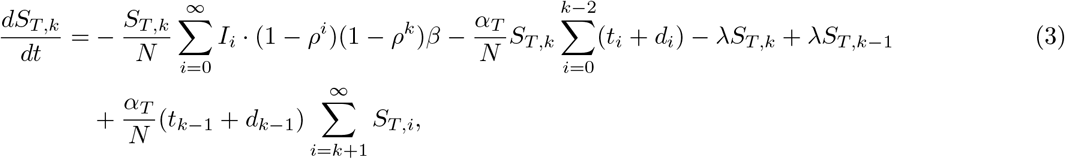

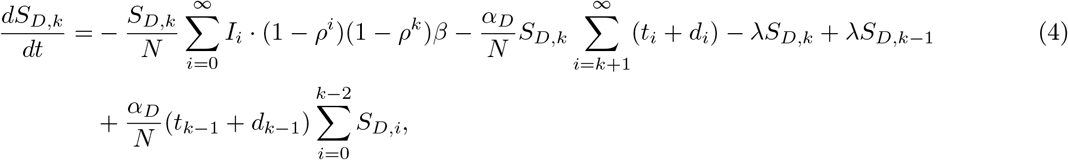

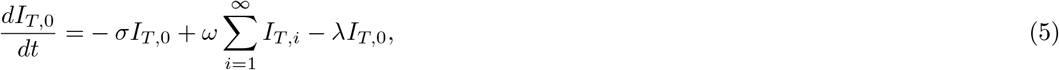

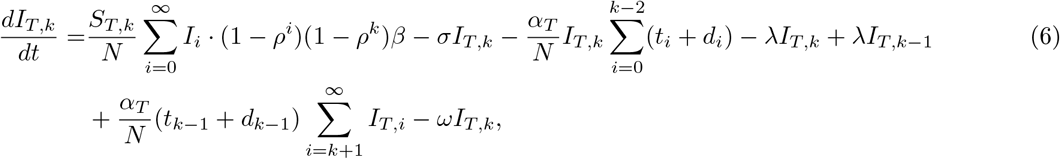

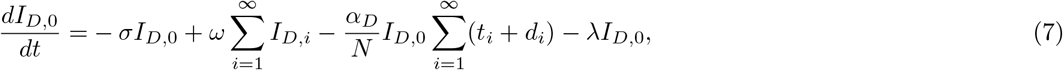

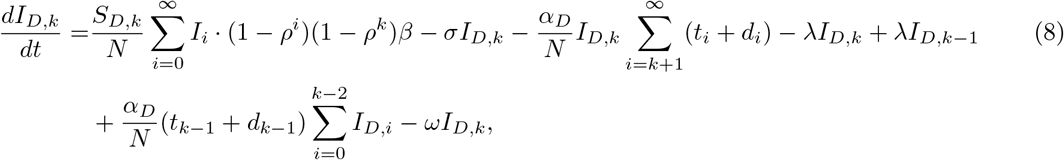

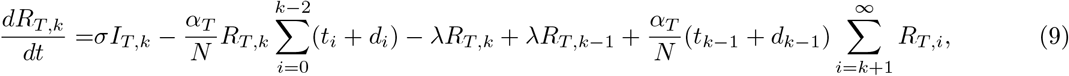

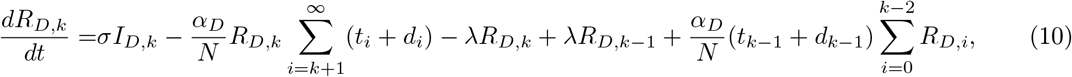

where *β* is the transmission rate in the absence of any protective measures, *σ* is the recovery rate, and *ρ* is a fixed parameter which measures the effectiveness of non-pharmaceutical interventions, 0 < *ρ* < 1. The first terms in Eqs. (3), (4), (6), and (8) correspond to new infections of susceptible individuals. In contrast with the model [13], we assume that disease transmission is reduced by both infected and susceptible individuals reducing their contact rates, hence the factor (1 − *ρ*^*i*^)(1 − *ρ*^*k*^) in the transmission rates. In this case, infectious individuals at information level 0 do not infect any susceptibles during the period they are fully aware of their condition. This is the case for diseases for which self-isolation is imposed after the detection of the disease - assuming the disease is detected before the individual recovers.

The second link between disease and awareness dynamics is obtained through the second terms in Eqs. (5) and (7) that describe the information generation by infected individuals. Information is generated when infected individuals realise their condition, for example following a positive test result. This effect distinguishes between diseases with unmistakable symptoms and cases where the infection is contagious but asymptomatic, or where the infection might be mistaken for another condition. The parameter *ω* then varies not only from illness to illness, but it also depends on the mechanisms to identify the disease quickly, such as fast testing and contact-tracing programmes.

Finally, the *σI*_*Y,j*_ terms in Eqs.(5–10) correspond to the recovery of infected individuals. The remaining terms correspond to the previously described information dynamics (see Eqs.(1–2)), giving the changes in awareness level due to information exchange and fading. Summing over the information subscripts leads to

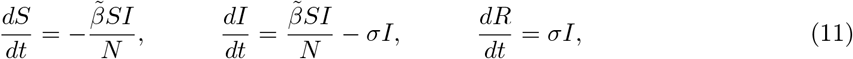

where

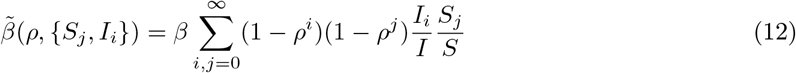

yields an effective transmission rate due to protective measures taken by informed individuals. Here, *I*_*i*_ = *I*_*C,i*_ + *I*_*D,i*_ and *S*_*j*_ = *S*_*C,j*_ + *S*_*D,j*_ as before, and *I* = ∑_*i*_ *I*_*i*_ and *S* = ∑_*i*_ *S*_*i*_. This is similar to the SIR model, with *β* varying as the epidemic progresses. In fact, our model reduces to the SIR model if no awareness is present in the population and awareness generation is turned off, i.e., the population is distributed within subpopulations *X*_*Y*,∞_, and *ω* = 0, respectively. In the SIR model, the epidemic threshold is at *R*_0_ = *β*/*σ* = 1 [23]. In our model, the effective reproduction number has a similar form, 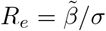. If we start with a fully uninformed and susceptible population, in which case awareness arises only through the process of information generation, then 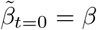, and the epidemic threshold is again at *R*_*e*_ = *β*/*σ*. Only if a certain level of awareness were already present in the population at the beginning of the epidemic, would the threshold be reduced to 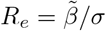, where 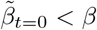.

Notice that no transmission would occur if the population was completely informed with the best quality of information. This situation does not arise, however, because susceptibles can at best obtain an index 1 status if they are informed by infected individuals with first-hand information, *I*_0_.

There is now a mutual feedback between information and disease dynamics, where information is produced by newly infected individuals, increasing the level of awareness in the trusting population and eliciting higher levels of protection through a reduction in contacts and adherence to other non-pharmaceutical interventions, hindering the spread of the disease. With sufficiently fast spread of information in the trusting population, leading to a quicker response to an increase in the number of infections, a fine balance can be achieved. The presence of distrusting individuals in the population reduces the effectiveness of the protection generated by the trusting population, threatening this delicate equilibrium. In the next section, we investigate the effects of varying the speed of information spread parameters, *α*_*T*_ and *α*_*D*_, the information-mediated protection parameter, *ρ*, and the density of distrusting individuals in the population, *d*, on the course and outcome of an epidemic.

## 3. Results

During an epidemic, the extent to which people comply with restrictions, such as mask-wearing, self-isolation, and social distancing, and the effectiveness of such protective measures is pivotal to control the spread of the disease [24]. In our model, the parameters that quantify these quantities are the density of distrusting individuals in the population, *d*, and the information-mediated protection parameter *ρ*.

### 3.1. The effects of varying *ρ* and *d*

Consider an initially unaware population composed of distrusting and trusting individuals with density *d* and 1 − *d*, respectively, where a small number of infected individuals are introduced in the population. We assume that the composition of trusting and distrusting populations remains constant during the course of the epidemic. Initially, there is no awareness in the population, and consequently no protection stemming from behavioural change. Hence, the disease spreads following a classic *SIR* model, where the threshold 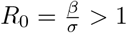 gives the condition for an initial increase in the number of cases.

As in the SIR model, if *R*_0_ < 1, the disease cannot spread in the population even in the absence of behavioural changes to control it. The awareness arising from the initial cases increases protection in the population and reduces the effective reproduction number *R*_*e*_ even further, speeding up the process of disease die out.

If *R*_0_ > 1, after an initial phase of unrestricted spread, the infected individuals will start to become aware of their condition, generating fresh information in the population. For a single infected individual, the time taken is of order *ω*^−1^. This awareness then spreads according to the information dynamics, eliciting protective measures among the informed individuals and causing a reduction in the effective reproduction number *R*_*e*_. If the response to new infections is sufficiently strong, a fine balance can be achieved, in which the increase in cases is counterbalanced by more cautious behaviour in the population. The presence of distrusting individuals, however, limits the effectiveness of the information-mediated protection in the population, as they will only comply with protective measures if directly affected by the disease.

In Figure 1(a) we show the trajectories of the total number of infected over time, *I*(*t*), for different *ρ* values, fixing the remaining parameters. Similarly, Figure 1(b) shows *I*(*t*) for different distrusting densities *d*. We have chosen *R*_0_ > 1 so that we see non-trivial infection dynamics. In both cases, we observe an initial increase in the number of infected individuals that is common to all curves. This corresponds to the unaware phase with *R*_0_ > 1. After this initial phase of unrestricted contagion, there is a change in the curves’ behaviour as the parameters vary. For small *ρ* and large *d* values, the elicited protection is not able to suppress the outbreak after the initial contagion phase, in which case we observe an effect similar to a reduction in *β* for the SIR model, where the peak of infection occurs later, and the epidemic duration extends. In contrast, for large *ρ* values and small densities *d*, the evoked protective measures taken by the informed individuals are sufficient to bring and keep *R*_*e*_ below 1. The epidemic outbreak is then suppressed, and we observe a peak of infection happening earlier in the epidemic, also reducing its duration.

**Figure 1.**
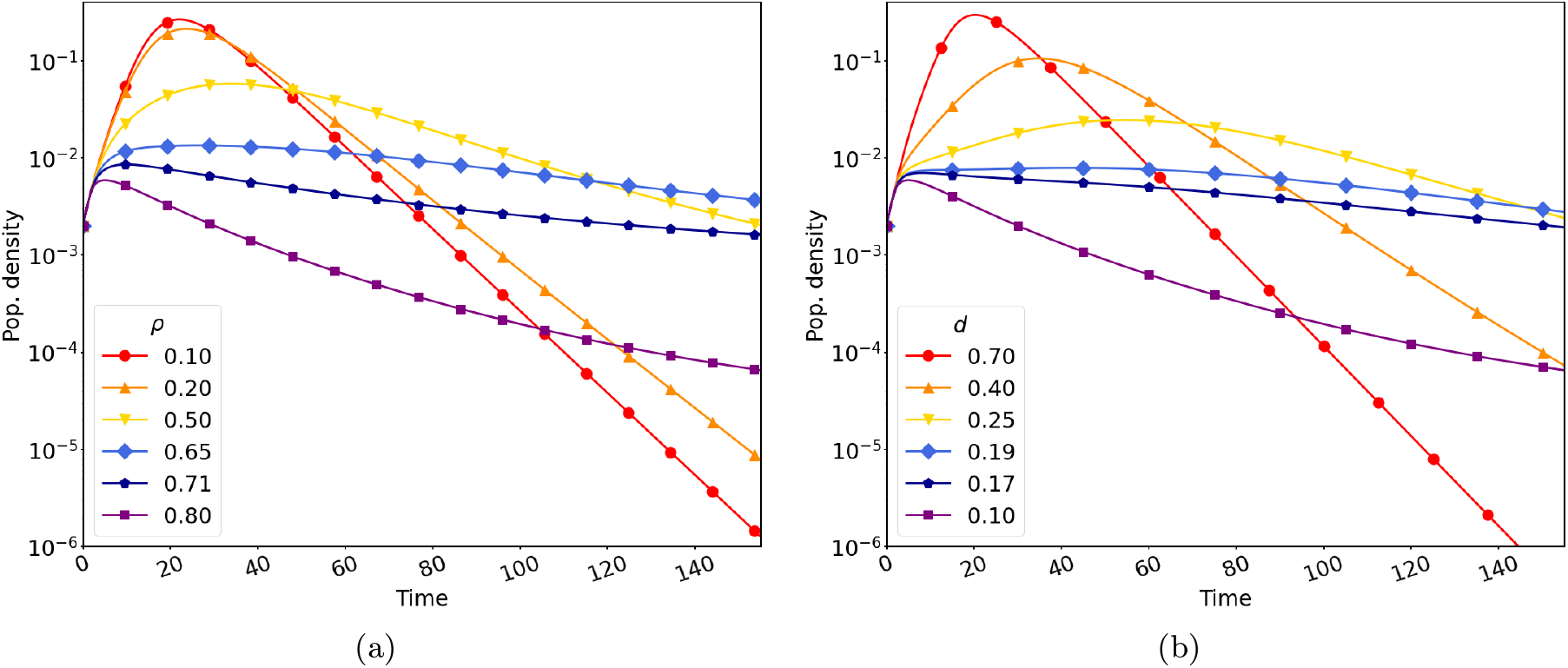
Stronger interventions (increasing *ρ*) have a similar effect on disease dynamics as a more trusting population (decreasing *d*). Curves show time series of the total number of infected *I*(*t*) for varying (a) *ρ* (fixed *d* = 0.1) and (b) *d* (fixed *ρ* = 0.8). In both cases *α*_*T*_ = *α*_*D*_ = 5, *β* = 0.667, *σ* = 0.133 (*R*_*e*_(0) = *R*_0_ = 5), *λ* = 0.2, and *ω* = 0.333.

For intermediate values, we observe the delicate balance between new infections, generating awareness, and the reduction in transmissibility by the informed individuals. This leads to a plateau in the number of infected individuals in the population and to really long epidemics. A similar effect has been observed by Weitz et al. [12] in the context of a SEIR model without awareness spread, but including awareness-driven behavioural changes. There are critical values *ρ*_*c*_ and *d*_*c*_ for which we observe a phase transition in the behaviour of the epidemic. For *ρ* < *ρ*_*c*_ (or *d* > *d*_*c*_), the epidemic spread is only mitigated, while for *ρ* > *ρ*_*c*_ (*d* < *d*_*c*_) the outbreak is suppressed by the behavioural changes in the population. The value of *ρ*_*c*_ depends on the density of distrusting individuals in the population, and similarly *d*_*c*_ depends on the information-mediated protection levels. We investigate this dependence, *ρ*_*c*_ = *ρ*_*c*_(*d*), further in Sec. 3.2.

In the real world, resources such as hospital staff, ventilators, and ICU beds are limited. Thus, knowing how many people will be affected at the same time and require medical assistance is essential to manage epidemics. Not only that, being aware of the factors that influence the size of the peak of infection is extremely valuable to aid policy-making. To explore further the effects of *ρ* and *d* on the peak of infection, we solved Eqs.(3–10) numerically for different values of *α* whilst again varying *ρ* and *d*.

Figure 2 shows the size of the peak of infection and the final susceptible population as a function of the parameters *ρ* and *d* for different *α*_*T*_ values. A common effect of behavioural feedback in epidemics is the reduction in the number of infected and increase in the final susceptible population [7, 12, 13]. We obtain a similar result in Figure 2(a) and 2(c). Increasing the information-mediated protection in the population, *ρ*, monotonically decreases the size of the peak of infection and hampers the spread of the disease, resulting in larger susceptible populations at the end of the epidemic.

**Figure 2.**
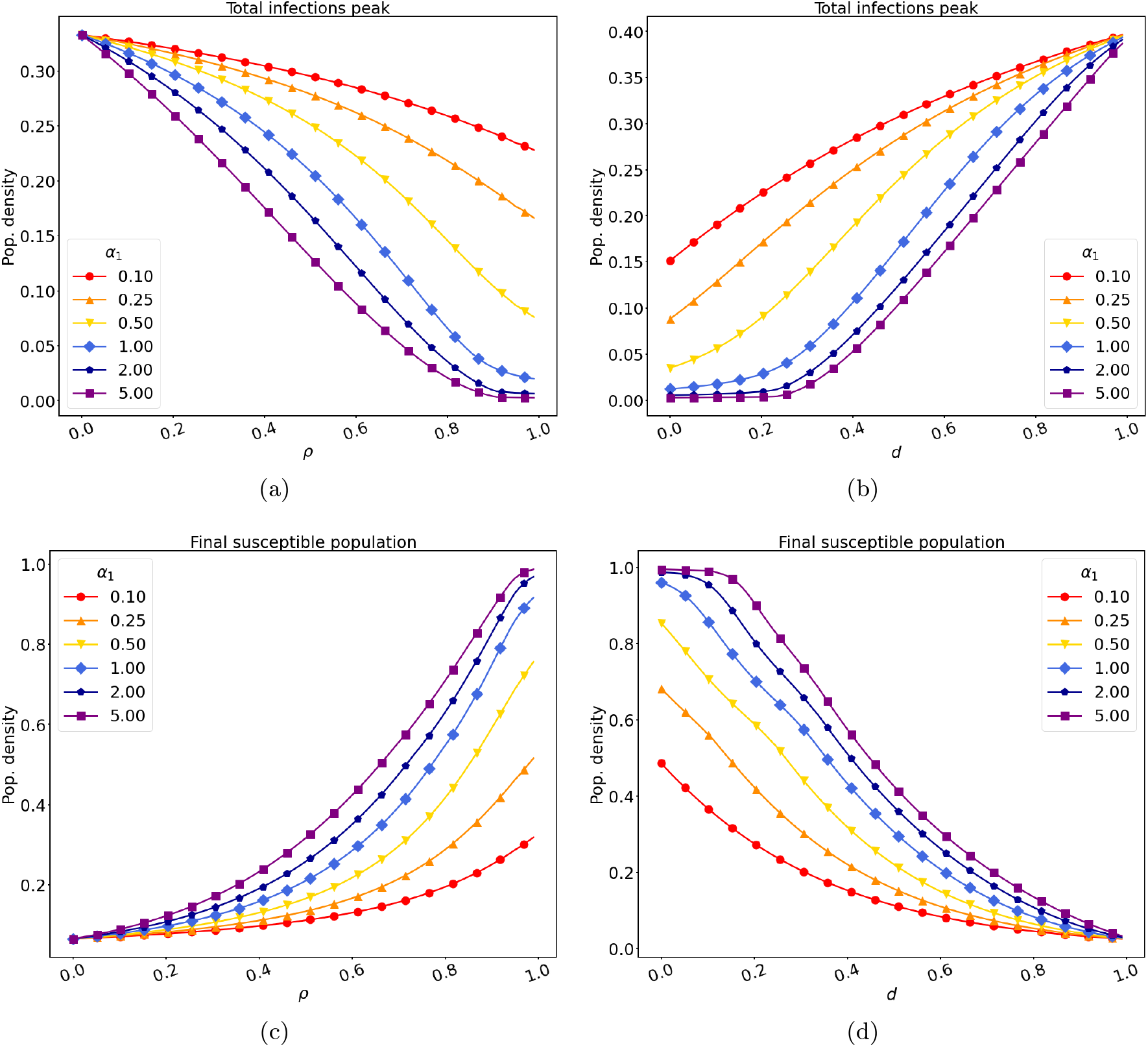
Faster information spread (increasing *α*) heightens model sensitivity to the strength of interventions (*ρ*) and composition of the population (*d*). Curves show effects of varying *ρ* and *d* on the peak infection size and the final susceptible population. (a) Total infections peak for varying *ρ* and fixed *d* = 0.3. (b) Total infections peak for fixed *ρ* = 0.8 and varying *d*. (c) Final susceptible population for varying *ρ* and fixed *d* = 0.3. (d) Final susceptible population for fixed *ρ* = 0.8 and varying *d*. Remaining parameters: *α*_*D*_ = 1, *λ* = 0.2, *ω* = 0.333, *β* = 0.667 and *σ* = 0.133.

In contrast, in Figures 2(b) and 2(d) we observe that larger densities of distrusting individuals monotonically increase the size of the peak of infection and lead to more overall infections, resulting in smaller susceptible populations at the end of the epidemic. This stems from the reluctance that this portion of the population has to abide by protective measures.

Our results highlight the importance of faster information spread in the trusting population, the use of non-pharmaceutical control measures, and the development of information campaigns to better inform the population and increase compliance with protective measures as a means to reduce disease spread and lighten the disease-associated healthcare burden.

Other important disease outbreak metrics are the expected duration of an epidemic and the time when the peak of infection occurs. Figure 1 already gave us an idea of how these quantities vary with *ρ* and *d*. To get a better picture of their effect, we solved the system of equations numerically again and measured the time until the peak of infection for different combinations of *ρ* and *d*, and different *α*_*T*_ values. The results can be seen in Figure 3. The initial condition consisted of an almost entirely susceptible, unaware population with initial infected population density 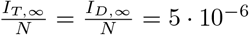.

**Figure 3.**
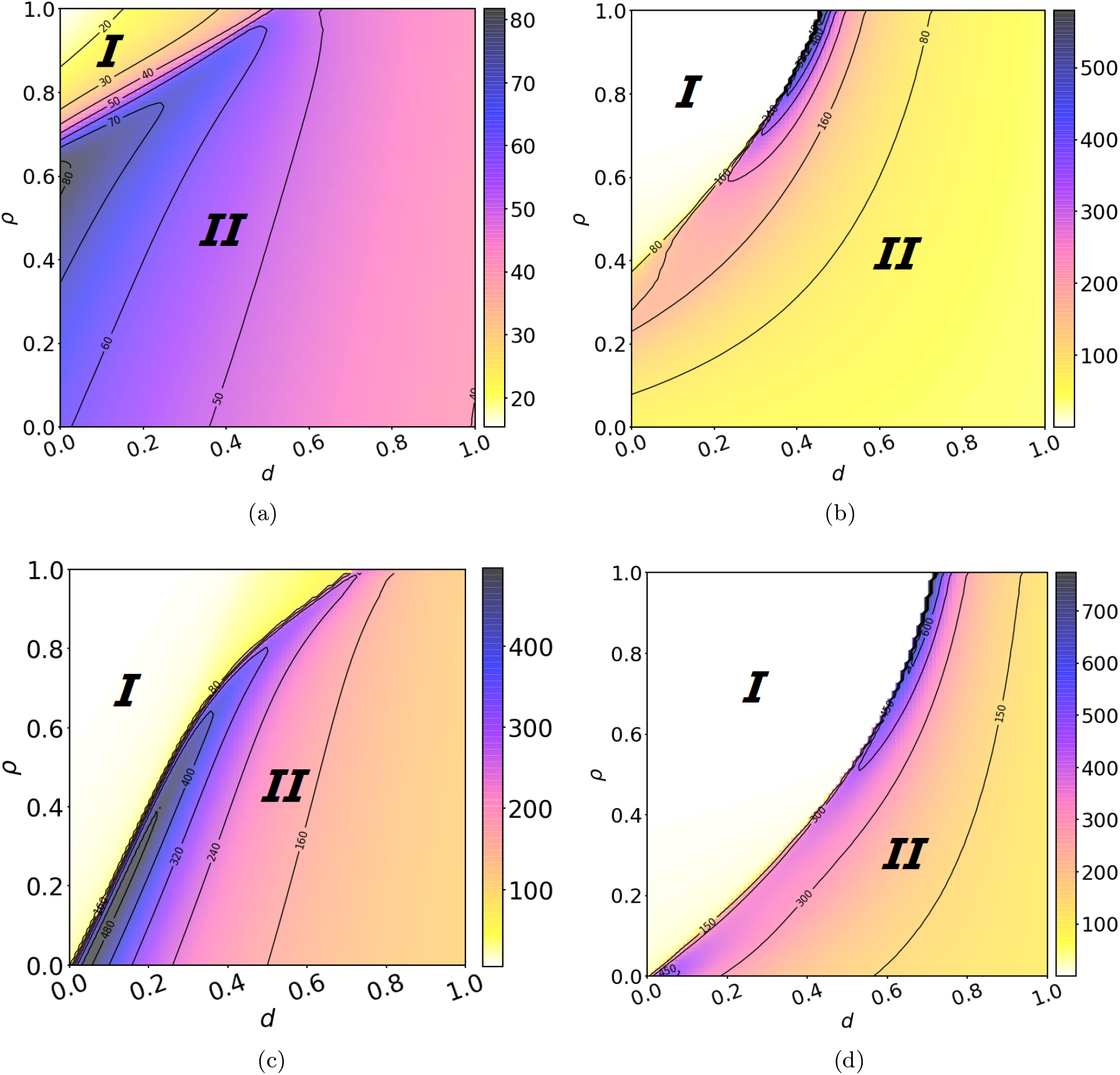
Timing of the infectious peak (colours) reveals the boundary between epidemic suppression and large outbreaks. First row: *R*_0_ = 3.5 (*β* = 0.467), (a) *α*_*T*_ = 0.5 and (b) *α*_*T*_ = 500. Second row: *R*_0_ = 2 (*β* = 0.267), (c) *α*_*T*_ = 0.5 and (d) *α*_*T*_ = 500. The remaining parameters are: *α*_*D*_ = 1, *λ* = 0.2, *ω* = 0.333 and *σ* = 0.133.

The first thing to notice from Figure 3 is the existence of the two regions mentioned earlier and the phase-transition, given by a curve *ρ*_*c*_(*d*). The first region (*I*), where *ρ* > *ρ*_*c*_(*d*), is the suppression regime, characterised by very low peaks of infection and times until the peak occurs that decrease as *ρ* increases. The second (*II*), where *ρ* < *ρ*_*c*_(*d*), is the mitigation regime, characterised by larger peaks of infection. In the mitigation regime, although the infection peak decreases as *ρ* increases, the times until the peak of infection increase, in contrast to the suppression regime. At the boundary, we observe a phase-transition, where the response to new infections is just strong enough to contain the increase in cases. At this boundary, we observe almost constant infection levels and, consequently, extended epidemics. The transition between the regions becomes more abrupt for larger *α*_*T*_ values, i.e., when awareness transmission is much faster in the trusting population.

In addition to the two regions, we note the existence of a critical value *d* = *d*_*M*_ above which suppression of the disease is not possible. This significant result highlights the need for effective measures to combat the spread of misinformation and the importance of maintaining trust in order to contain and suppress the disease. Furthermore, increasing the speed of information spread in the trusting population reduces *ρ*_*c*_(*d*), increasing the area of parameter space for which suppression is achievable, in contrast with the increase of *α*_*D*_, which increases *ρ*_*c*_(*d*), reducing the suppression region of the parameter space.

In essence, we observe a second threshold in the model. The first one corresponds to the outbreak threshold, where the disease can initially spread in the unaware population, given by *R*_0_ > 1. The second corresponds to the suppression threshold, where the protection elicited in the trusting population by the initial infection can reduce *R*_*e*_ to less than 1, hampering the spread of the disease. A second threshold due to preventive behaviour has been seen in the context of a SIS-like epidemiological model with awareness feedback on regular random networks and absence of fading mechanism [25], leading to a slow die-out of the epidemic and to a final population with a significant number of aware individuals. It has also been observed in [15, 16] in the case of a SIRS model with two states of aware and unaware individuals. In [15, 16], the authors observe that awareness changes the invasion conditions between a disease-free and endemic equilibrium, and can make it impossible for a disease to establish itself in the population. In both cases, the endemic equilibriums are characteristic of the chosen model, and the threshold can only be observed if awareness does not deteriorate through transmission or fading.

Naturally, the second threshold depends on the characteristics of the information spread dynamics, *α*_*T*_ and *α*_*D*_, the link between the effectiveness of behavioural change *ρ*, and the population composition, *d*. This threshold can be reflected in the effectiveness of protection elicited by behaviour change *ρ*_*c*_ given the population composition (density of distrusting individuals, *d*), and a fixed speed of information spread *α*_*T*_ and *α*_*D*_. Smaller *ρ*_*c*_ values mean that less protective measures must be taken to reach the suppression threshold.

In the next section, we derive a theoretical expression for the value of *ρ*_*c*_ as function of *d* in the limit of fast information spread, i.e., large *α*_*T*_ and *α*_*D*_. In this case, we observe a separation of timescales between disease and information spread, allowing an analytical treatment of the simplified model.

### 3.2. Estimates of the critical *ρ* and density values under fast spread of information

Although it is difficult to express the suppression threshold analytically in most cases, as it depends on the distribution of awareness in the population and its complicated dynamics, we can make use of extra assumptions, such as fast information spread and large population size *N*, to simplify the model. The procedure is similar to obtaining the basic reproduction number in most epidemiological models [26].

Consider an initially unaware population of size *N* with a small density of infected individuals. Additionally, consider the limit in which the spread of information happens almost instantaneously, i.e. *α*_*T*_ → ∞ and *α*_*D*_ → ∞. This would be the case, for instance, where new cases are informed globally through the internet or mass media as soon as they are discovered. In this limit, there is a separation of time-scales between the information and disease spread dynamics. A brief moment after the beginning of the epidemic, the first infected individual refreshes their information, moving to index 0. Immediately, all remaining trusting individuals in the higher index levels move to index 1, while all distrusting individuals remain unaware. Note that once infected distrusting individuals refresh information, they immediately become unaware again. We can then assume that the distrusting population occupies only the *S*_*D*,∞_, *I*_*D*,∞_ and *R*_*D*,∞_ subpopulations. In this scenario, the model simplifies to

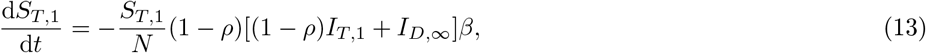

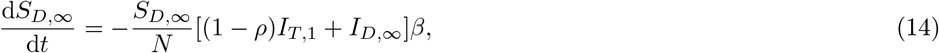

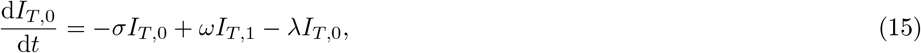

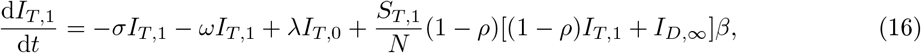

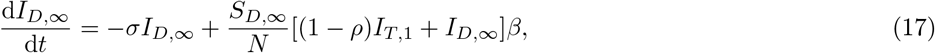

where we have omitted the equations for the recovered populations. We drop the indices *T* and *D* for convenience as there is no shared information index between the two groups. As observed in the result section, we assume that the level of infected individuals is approximately constant (metastable) at the phase transition

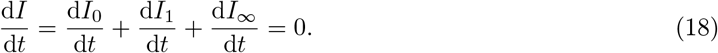

We also assume metastability of all three infected groups. Setting 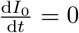 gives 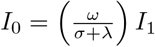. Setting Eq. (17) to zero, and assuming a large population such that, after the initial phase of unrestricted transmission, 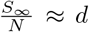, where *d* is the initial density of distrusting, and 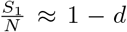, results in 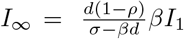. Now, from Eq. (18),

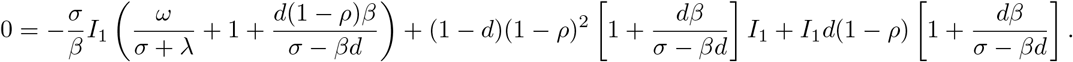

Denoting *x* = (1 − *ρ*) and 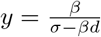,

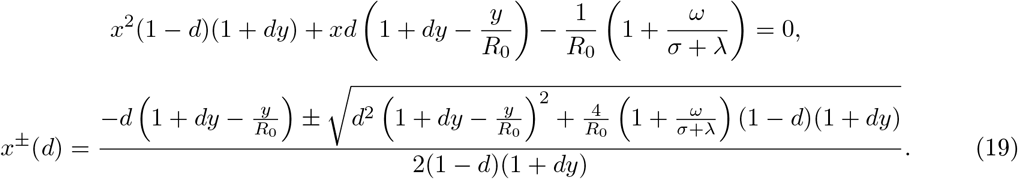

We then obtain the critical protection level *ρ*_*c*_ as a function of the distrusting population density *d, ρ*_*c*_(*d*) = 1 − *x*(*d*). For *ρ* < *ρ*_*c*_(*d*) we observe mitigation of the disease spread, whilst for *ρ* > *ρ*_*c*_(*d*) the disease is suppressed.

From the expression above, we note that there is a critical distrusting density 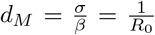 above which suppression of the disease is not feasible irrespective of the level of information-mediated protection, *ρ*. Solutions for *d* > *d*_*M*_ yield negative *I*_*∞*_ population sizes and are, therefore, not physical. Similarly, we are interested in solutions for which 0 < *ρ* < 1, or equivalently, 0 < *x* < 1. Notice that, for *d* < *d*_*M*_, the *x*^+^ solution is always greater than zero, giving *ρ*_*c*_ < 1. Hence, there always exists a region for *d* < *d*_*M*_ in which suppression of the disease is achievable for sufficiently large *ρ*. Notice that, if *R*_0_ < 1, then *d*_*M*_ > 1. Hence, suppression is always achieved in this case. This is consistent with the threshold for an initial outbreak *R*_0_ > 1.

In particular, for *d* = 0,

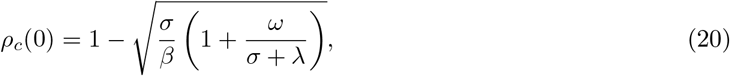

gives us conditions where *ρ*_*c*_(0) > 0 and we can see a phase transition from mitigation to suppression for increasing *ρ* in the absence of distrusting individuals. If *ρ*_*c*_(0) < 0, then suppression is achieved for all *ρ* values, and we do not observe the transition as *ρ* increases. From Eq. (19), and more easily from Eq. (20), we can see that increasing *ω* while keeping the remaining parameters constant reduces *ρ*_*c*_(*d*), since more information is being produced and transmitted, which increases awareness in the population. In contrast, increasing *λ* increases *ρ*_*c*_(*d*) as the information is forgotten faster. Similarly, increasing *σ* while keeping *R*_0_ constant increases *ρ*_*c*_(*d*), as the probability that individuals recover before realising they were infected increases.

Figure 4 shows the regions of *d* − *ρ* parameter space corresponding to suppression (*I*) and mitigation (*II*) obtained through the numerical solution of the complete model (with large *α* and *I*_*T,∞*_/*N* = *I*_*D,∞*_/*N* = 5 · 10^−6^), and the theoretically predicted boundary (dashed black line), Eq. (19), for two values of *β* (and corresponding two values of *R*_0_). Good agreement was also observed for different combinations of *β* and *σ* (not shown), although discrepancies arise as the rate of information spread is reduced. Nonetheless, Eq.(19) shows the existence of regions of suppression and mitigation and a critical density of distrusting individuals above which suppression is not achievable, emphasising the importance of effective measures to combat misinformation and increase the population’s compliance with disease-control measures.

**Figure 4.**
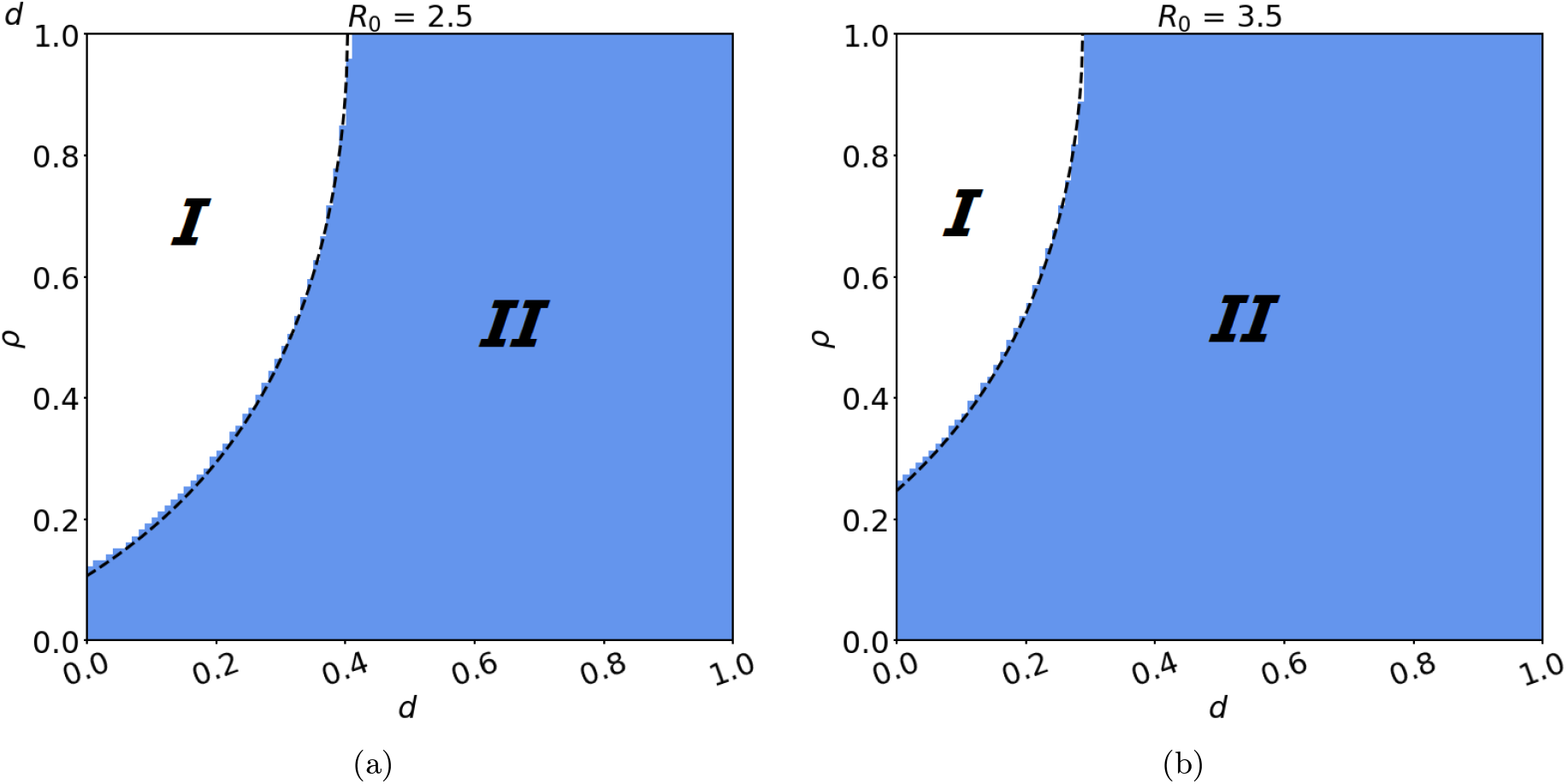
The agreement between the theoretical prediction (dashed lines) of the observed phase transition and the suppression/mitigation regions obtained through the numerical solutions of Eqs.(3–10) for *α*_*T*_ = *α*_*D*_ = 10^6^, *λ* = 0.2, *ω* = 0.33, *σ* = 0.133, and (a) *β* = 0.333 (*R*_0_ = 2.5) and *β* = 0.467 (*R*_0_ = 3.5). The white area (*I*) correspond to the region where disease spread is suppressed. The area in blue (*II*) correspond to the region where disease spread is only mitigated by behavioural feedback.

## 4. Discussion

Mathematical models are pivotal to policy-making and managing epidemics, giving insight into important aspects of the dynamics of a disease that can facilitate its control and predict possible outcomes. In the past, epidemiological models failed to account for the important aspect of behavioural responses to diseases in human populations. More recently, prominent approaches focusing on the spread of awareness and its interplay with disease dynamics have been developed [5].

In this article, we developed a model that merged epidemiological dynamics with information spread and behavioural change interactions of both trusting and distrusting individuals in a population. This model is an extension of previous work in this area which considered only one class of individuals [13]. Using our model, we were able to highlight the mechanisms by which the burden of epidemics can be lightened.

Our results show that the surge in awareness, arising from fast spread of information in the trusting population, can reduce the peak of infection and lead to a larger population that remains uninfected by the end of the outbreak. A similar effect is observed by increasing the effectiveness of protective measures taken by well-informed individuals. We also observed a phase transition between mitigation and suppression regimes. In the mitigation regime, the behavioural reaction and reduction in transmissibility arising in the population is not sufficient to reduce the effective reproduction number below one. Hence, the observed effect in the infection curve is a lower and delayed peak. However, if the response to new cases is sufficiently strong, a fine balance between new infections and a reduction in transmissibility can be achieved. In this case, we observe an almost constant level of infection over time, leading to extended epidemics. We then observe a phase transition, in which further increasing the effectiveness of the measures to control the spread of the disease leads to a regime of suppression. In contrast, larger densities of distrusting individuals in the population limit the effectiveness of the reduced transmissibility in the trusting population, resulting in the existence of a critical density *d*_*M*_ from which the suppression regime is not achievable.

Although a complete and simple analytical characterisation of the suppression threshold is difficult in general, as it depends on the complex dynamics of the system, we were able to write an expression for *ρ*_*c*_(*d*) in the limit of fast information transmission and assuming a large population size. The expression agreed with the numerically observed boundary in the corresponding parameter regime, and allowed us to obtain the maximum proportion of distrusting individuals for which suppression of the outbreak is achievable. A possibly fruitful direction for further research to obtain approximations for realistic information transmission parameter values, but that still exhibit slow and fast dynamics, is the analysis of the stochastic version of the system using the framework developed in [27] that allows for dimension reduction in stochastic dynamical systems with a separation of timescales. The method is exact in the limit of well-separated fast-and-slow dynamics and small noise, and found to be a reasonable approximation over a sensible parameter range.

The deterministic model developed has the underlying assumption of a well-mixed population, for which the explicit spatial structure of the system is disregarded. Previous work on the effects of spatial structure in the spread of infectious diseases has shown its importance in the invasion threshold for epidemics [28], and the development of control policies [29–32]. The analysis of epidemiological models with awareness feedback on networks with homogeneous populations has been an active area of research [10, 33–38]. However, these models disregard the heterogeneous behaviour presented here. Thus, the inclusion of spatial dynamics in our model is another prospective direction for further work.

## Data Availability

Not applicable.

## Acknowledgements

A.S. is supported by a scholarship from the EPSRC Centre for Doctoral Training in Statistical Applied Mathematics at Bath (SAMBa), under the project EP/S022945/1. T.R. acknowledges the support of the Royal Society RGF\EA\180242. This research made use of the Balena High Performance Computing (HPC) Service at the University of Bath.

## Notes

### Competing Interest Statement

The authors have declared no competing interest.

